# Treatment Efficacy of Theophylline in ADYC5 Dyskinesia: A Retrospective Case Series Study

**DOI:** 10.1101/2024.08.23.24312408

**Authors:** Dirk Taenzler, Frank Hause, Andreas Merkenschlager, Andrea Sinz

**Author notes:** **Corresponding author:** Prof. Dr. Andrea Sinz, Center for Structural Mass Spectrometry, Martin Luther University Halle-Wittenberg, 06120 Halle (Saale), Germany.

## Abstract

**Background:** ADCY5-related dyskinesia is a rare disorder caused by mutations in the ADCY5 gene resulting in abnormal involuntary movements. Currently, there are no standardized guidelines to treat this condition.

**Objectives:** The aim of this study is to evaluate the efficacy of theophylline administration in improving symptoms and quality of life in patients with ADCY5-related dyskinesia.

**Methods:** A retrospective study was conducted involving 12 patients (aged 2-34 years) with ADCY5-related dyskinesia. Participants completed a questionnaire about theophylline administration, including dosage, improvement of symptoms, adverse effects, and changes in quality of life. Data were analyzed for reported efficacy and side effects.

**Results:** Theophylline administration demonstrated substantial efficacy, with 92% (11 out of 12) of patients reporting significant improvements in their movement disorders. The average improvement score was 7.0 (± 1.9) on a 10-point scale. Notable improvements included reductions in severity and frequency of episodes, improved gait, more independent mobility, psycho-social well-being, and quality of sleep. Adverse effects were reported by 6 patients, including dystonia, speech worsening, headaches, nausea, impaired sleep, and agitation.

**Conclusions:** Theophylline shows substantial promise as a treatment option for ADCY5-related dyskinesia, improving various aspects of patients’ quality of life and movement disorder symptoms. Further research is needed to optimize dosing, to understand long-term effects, and to explore combinational drug therapies. Despite the small cohort size and the retrospective nature of this study, the results support theophylline administration to decrease dyskinetic movements and enhance overall quality of life in patients.

## Introduction

ADCY5-related dyskinesia is a rare neurological condition caused by mutations in the ADCY5 gene that codes for the enzyme adenylyl cyclase 5. This enzyme is essential for intracellular signaling, as it converts ATP into cyclic AMP (cAMP), a secondary messenger involved in numerous cellular functions. Mutations in the ADCY5 gene cause increased enzymatic activity,^1^ leading to an overproduction of cAMP and clinically resulting in abnormal involuntary movements referred to as dyskinesia.^2^

ADCY5-related dyskinesia manifests through a variety of impaired voluntary movements, including but not limited to choreoathetosis, dystonia, myoclonus and/or axial hypotonia,^3^ ultimately resulting in poor motor control. These symptoms typically emerge in infancy or early childhood, affecting the limbs, face, and neck.^4^ The movements are often continuous during waking hours and can disrupt sleep. They tend to worsen with stress, fatigue, or illness but may also occur without any identifiable trigger.^4^ The majority of individuals diagnosed with ADCY5-related dyskinesia are simplex cases, meaning they are the only affected member in their family, and their condition arises from a de novo pathogenic variant. Diagnosis is primarily based on clinical presentation and confirmed through genetic testing to identify mutations in the ADCY5 gene.^5-7^

Based on case reports, patients with ADCY5-related dyskinesia may benefit from deep brain stimulation (DBS).^8-10^ Bilateral pallidal DBS can significantly reduce hyperkinetic movements and paroxysomal episodes. While there is generally partial improvement patients often remain impaired and daily living activities may not significantly improve.^11^

Currently, there are no randomized controlled trials or consensus guidelines for treating ADCY5-related dyskinesia. Case reports suggest that benzodiazepines, such as clonazepam, clobazam, diazepam, and lorazepam, can help managing dyskinetic episodes, also those associated with sleep.^8^ In a retrospective study involving 30 individuals, a beneficial effect of caffeine on hyperkinetic movements was observed.^12^ This effect is thought to be attributed to the attenuating action of caffeine as an adenosine 2A antagonist, which is believed to lead to a reduction of cAMP levels in the striato-pallidal projection neurons.^12^

We were able to demonstrate that treatment with the purine derivatives caffeine, theophylline, and istradefylline reduced ADCY5-catalyzed cAMP production in cell lines overexpressing wild-type and mutant ADCY5. The most pronounced effects on cAMP reduction were observed in ADCY5 R418W mutant cells^13^, a mutation found in the majority of individuals with ADCY5-related dyskinesia^14^. Following these findings, a slow-release theophylline formulation was administered to a preschool-aged patient with ADCY5-related dyskinesia, resulting in a striking improvement of symptoms, surpassing the effects of caffeine that had previously been administered to the same patient.^13^

Based on these encouraging results, we systematically present the first comprehensive account of the effects of theophylline administration in a series of retrospective reports from patients with ADCY5-related dyskinesia.

### Patients and Methods

Patients and/or their parents, if the patients were too young or unable to complete the questionnaire themselves, were asked to fill out a form that had been specifically designed for this study. The questionnaire targeted individuals who had administered a slow-release formulation of theophylline to treat ADCY5-related dyskinesia. The questionnaire was made available to interested participants via the patient organization portal adcy5.org. It was provided in German, English, French, and Spanish. Interested individuals were given the opportunity to answer the questionnaire anonymously. The questionnaire was uploaded by DT, and the collection of responses including their anonymization was also conducted by DT. Anonymized forms were forwarded to AM for analysis.

The aim of the ADCY5 questionnaire for parents was to systematically gather comprehensive information about symptoms, treatment responses, and overall patients’ quality of life with ADCY5-related movement disorders and potential changes following theophylline administration. In addition to demographic questions, the questionnaire includes items related to the patients’ genotype, cardinal dyskinetic symptoms and their frequency, current theophylline dosage, positive and adverse effects of theophylline administration, as well as former and ongoing (co-)medications for the treatment of ADCY5-related dyskinesia.

The study protocol was approved by the Ethics Committee of the Medical Faculty, University of Leipzig, on April 30, 2024 and listed in the German Clinical Trials Register (DRKS00034720), on Juli 25, 2024. All patients, or their authorized representatives, provided written informed consent prior to participation in this study. Data produced in the present study are available upon reasonable request to the authors. The questionnaire provided to the patients is available as online supplemental data.

## Results

### Demographics

The study included 12 patients, comprising 7 females and 5 males. The patients’ age ranged from 2 to 34 years, with a median age of 8 years. All patients were characterized by ADYC5-related dyskinesia with a diverse range of baseline hyperkinetic and paroxysmal movement disorders. Genotyping data of the ADCY5 gene was provided by the 12 participants in the questionnaire and confirmed mutations in the ADCY5 gene, with R418W being the most prominent mutation, present in 9 out of 12 individuals.

### Theophylline Efficacy

Theophylline administration demonstrated substantial efficacy across the study cohort. Out of the 12 patients, 11 reported improvements in their movement disorder symptoms, constituting 92% of the cohort. The average improvement score on a 10-point scale was approximately 7.0, with a standard deviation of 1.9, indicating a high level of efficacy with minor variability.

These improvements were comprehensive, affecting various aspects of the disorder symptoms. Most patients noted a reduction in the severity and frequency of their episodes (8 of 12), with a substantial number reporting reduced duration per episode (8 of 12) (see Table 1).

**Table 1:**
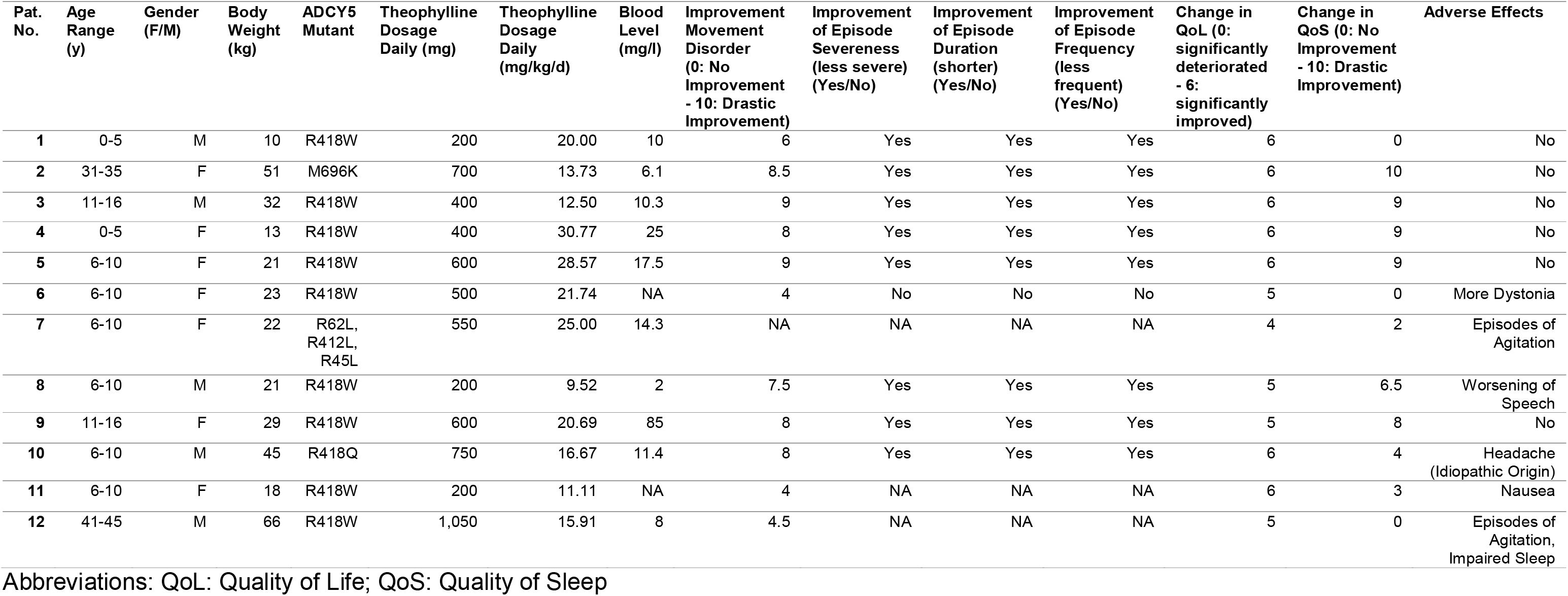
Characteristics of the patients and theophylline response.

| Pat. No. | Age Range (y) | Gender (F/M) | Body Weight (kg) | ADCY5 Mutant | Theophylline Dosage Daily (mg) | Theophylline Dosage Daily (mg/kg/d) | Blood Level (mg/l) | Improvement Movement Disorder (0: No Improvement - 10: Drastic Improvement) | Improvement of Episode Severeness (less severe) (Yes/No) | Improvement of Episode Duration (shorter) (Yes/No) | Improvement of Episode Frequency (less frequent) (Yes/No) | Change in QoL (0: significantly deteriorated - 6: significantly improved) | Change in QoS (0: No Improvement - 10: Drastic Improvement) | Adverse Effects |
| --- | --- | --- | --- | --- | --- | --- | --- | --- | --- | --- | --- | --- | --- | --- |
| 1 | 0-5 | M | 10 | R418W | 200 | 20.00 | 10 | 6 | Yes | Yes | Yes | 6 | 0 | No |
| 2 | 31-35 | F | 51 | M696K | 700 | 13.73 | 6.1 | 8.5 | Yes | Yes | Yes | 6 | 10 | No |
| 3 | 11-16 | M | 32 | R418W | 400 | 12.50 | 10.3 | 9 | Yes | Yes | Yes | 6 | 9 | No |
| 4 | 0-5 | F | 13 | R418W | 400 | 30.77 | 25 | 8 | Yes | Yes | Yes | 6 | 9 | No |
| 5 | 6-10 | F | 21 | R418W | 600 | 28.57 | 17.5 | 9 | Yes | Yes | Yes | 6 | 9 | No |
| 6 | 6-10 | F | 23 | R418W | 500 | 21.74 | NA | 4 | No | No | No | 5 | 0 | More Dystonia |
| 7 | 6-10 | F | 22 | R62L, R412L, R45L | 550 | 25.00 | 14.3 | NA | NA | NA | NA | 4 | 2 | Episodes of Agitation |
| 8 | 6-10 | M | 21 | R418W | 200 | 9.52 | 2 | 7.5 | Yes | Yes | Yes | 5 | 6.5 | Worsening of Speech |
| 9 | 11-16 | F | 29 | R418W | 600 | 20.69 | 85 | 8 | Yes | Yes | Yes | 5 | 8 | No |
| 10 | 6-10 | M | 45 | R418Q | 750 | 16.67 | 11.4 | 8 | Yes | Yes | Yes | 6 | 4 | Headache (Idiopathic Origin) |
| 11 | 6-10 | F | 18 | R418W | 200 | 11.11 | NA | 4 | NA | NA | NA | 6 | 3 | Nausea |
| 12 | 41-45 | M | 66 | R418W | 1,050 | 15.91 | 8 | 4.5 | NA | NA | NA | 5 | 0 | Episodes of Agitation, Impaired Sleep |

Abbreviations: QoL: Quality of Life; QoS: Quality of Sleep

Additional patient-reported outcomes include improvements in gait, more independent mobility, and increased psycho-social well-being in terms of confidence and happiness, as well as improved quality of sleep (on a 10-point scale: M = 5.0, SD = 3.8), significantly contributing to the overall improved functionality and quality of life reported by the patients.

### Theophylline Dosing and Tolerance

Theophylline was administered as a slow-release formulation at a daily dose ranging between 200 mg and 1,050 mg, adjusted to the patient’s body weight (9.5 to 28.6 mg/kg/day). The medication was generally well-tolerated. Adverse effects were reported by 6 patients. These included pronounced dystonia (1 of 6), worsening of speech (1 of 6), headaches of idiopathic origin (1 of 6), nausea (1 of 6), impaired sleep (1 of 6), and episodes of agitation (2 of 6).

### Other Treatments

Prior to theophylline administration, 10 patients had been treated with different regimens, including caffeine, levetiracetam, and benzodiazepines, with mixed results. After starting theophylline administration, 8 patients continued to use adjunct therapies, such as zopiclone and duloxetine, which were generally well-tolerated and enhanced overall treatment efficacy.

## Discussion

The present study indicates that theophylline administration substantially improves the symptoms and quality of life in patients with ADCY5-related dyskinesia, as evidenced by the high level of reported efficacy and alleviated symptoms.

Theophylline administration demonstrated substantial efficacy, with 92% of patients reporting improvements in their movement disorder symptoms. The average improvement score was 7.0 on a 10-point scale, with a standard deviation of 1.9, indicating a high overall efficacy. Improvements were noted in various symptom domains, including the severity and frequency of episodes, with 8 out of 12 patients reporting shorter episodes. Additionally, patients experienced improvements in gait, more independent mobility, enhanced psycho-social well-being, and higher quality of sleep, significantly contributing to their overall functionality and quality of life. Only minor adverse effects have been reported.

We are aware that there are several aspects in this study that should be considered. Firstly, the retrospective nature of the study introduces a potential bias and limits the ability to draw causal inferences. It is uncertain whether the participants were aware of the results from the initial case report involving the preschool-aged patient.^13^ Patients #2, #6, and #12 (see Table 1) had just started theophylline administration when completing the questionnaire. The relatively small cohort size is attributed to the rarity of ADCY5-related dyskinesia, which poses challenges to the feasibility of conducting randomized trials.

It has to be considered that the administration of theophylline might not improve sleep quality in ADCY5-related dyskinesia: Three patients (#1, #6, and #12, see Table 1) reported no improvement in sleep quality after theophylline administration. For patient #1, theophylline administration began at an age before the typical symptoms of the disease manifested. As evident from the overall improvement in quality of life for patient #1, theophylline has a positive effect, but this patient never experienced sleep disruption due to the disease. Patient #6 underwent surgery for DBS before administering theophylline, therefore it is difficult to attribute the effects to the theophylline treatment alone. Finally, patient #12 indicated that the reduced sleep quality might also be credited to the concurrent use of diazepam.

In conclusion, while theophylline demonstrates substantial promise as a treatment for ADCY5-related dyskinesia, further research is warranted to optimize dosing, to better understand long-term effects, and to explore the benefits of combinational drug therapies, especially regarding above mentioned limitations. Based on our results, we recommend theophylline administration in patients with ADCY5-related dyskinesia. Blood drug levels should optimally range between 15 to 20 mg/l to decrease dyskinetic movements and to improve overall quality of life.

## Supporting information

Questionnaire for Patients

## Data Availability

All data produced in the present study are available upon reasonable request to the authors. The questionnaire provided to the patients is available as online supplemental data.

## Authors’ Roles

DT, AM, and AS designed the study. DT informed willing participants about, and collected and anonymized the answers to the questionnaire. DT, AM, and FH analyzed the data. DT, FH, and AS wrote the final manuscript.

