## Supplementary material for "Treatment Efficacy of Theophylline in ADYC5 Dyskinesia: A Retrospective Case Series Study": Questionnaire for Patients

**ADCY5 Questionnaire for Parents**

*Patient Information:*

Gender:

Age:

Body weight:

Mutation site(s) in ADCY5:

Mosaicism: yes/no

1) *What are your child's symptoms before starting with theophylline treatment? Which ones are permanent, which ones occur by episodes (several days) or attacks (several minutes to a few hours)?*
   *Please rate the strength of symptoms with*
   *– (symptoms non-existent), x (weak symptoms), xx (strong symptoms), xxx (very strong symptoms)*

| | Permanent | Episode | Attack |
|---|---|---|---|
| *Dystonia (= muscle contractions resulting in twisting and repetitive movements)* | | | |
| *Myoclonus (= quick jerking movement of mime and gestures, especially in the region of the mouth that you cannot control)* | | | |
| *Chorea (= abnormal movements of the limbs)* | | | |
| *Tremor (= slight shaking movement)* | | | |
| *Gait / balance disorders* | | | |
| *Cramps* | | | |
| *Pain* | | | |
| *Attention and concentration deficits* | | | |

| |
|---|
| *Speech* |
| *Hypersalivation* |
| *Foot malpositioning* |
| *Core stability* |
| *Others (please specify)* |

2) *If your child has episodes (several days) or attacks (from several minutes to a few hours) what is their type, duration, and frequency? Were there "movement storms" at time of diagnosis* ***before*** *theophylline treatment?*

3) *On a scale of 0 (no improvement) to 10 (major improvement with total disappearance of symptoms), how do you rate the improvement with* ***theophylline*** *on your child's involuntary movements?*

0 1 2 3 4 5 6 7 8 9 10

4) *At which dose does your child take theophylline? Since when does he/she take his/her theophylline at this dose (morning- lunch- evening)? Does your child take a slow-release form of theophylline?*

5) *What is the current blood level (mg/l) of theophylline?*

*6) Since taking theophylline, would you say that your child's quality of life is:*

*- much improved*

**-** *improved*

*- minimally improved*

*- neither improved nor worse*

*- minimally worse*

*- much worse*

*- very much worse*

*7) Which symptoms are improved under theophylline treatment?*

*– (symptoms not improved), + (symptoms somewhat improved), ++ (symptoms improved), +++ (symptoms much improved)*

| | Permanent | Episode | Attack |
|---|---|---|---|
| *Dystonia (= muscle contractions resulting in twisting and repetitive movements)* | | | |
| *Myoclonus (= quick jerking movement of mime and gestures especially in the region of the mouth that you cannot control)* | | | |
| *Chorea (= abnormal movements of the limbs)* | | | |
| *Tremor (= slight shaking movement)* | | | |
| *Gait / balance disorders* | | | |
| *Cramps* | | | |
| *Pain* | | | |
| *Attention and concentration deficit* | | | |
| *Speech* | | | |
| *Hypersalivation* | | | |

| |
|---|
| *Foot malpositioning* |
| *Core stability* |
| *Others (please specify)* |

8*) a. If your patient has episodes or attacks with theophylline are they:*

*- less severe: yes / no*

*- shorter: yes / no*

*- less frequent: yes / no*

*b. If so, what was/is the duration of episodes (several days) or attacks (several minutes to a few hours) and how often did/do they happen (frequency) before and after starting theophylline treatment?*

*- episode frequency before theophylline:*

*- episode frequency after theophylline:*

*- episode duration before theophylline:*

*- episode duration after theophylline:*

*- attack frequency before theophylline:*

*- attack frequency after theophylline:*

*- attack duration before theophylline:*

*- attack duration after theophylline:*

*9) How do you evaluate the quality of sleep after theophylline treatment? Please rate the quality of sleep from 0 (no improvement) to 10 (major improvement).*

0 1 2 3 4 5 6 7 8 9 10

*10) Has your child presented any other positive effects in addition to those we talked about with theophylline, if so which ones?*

*11) What is the best improvement you can see?*

*12) Has your child had negative effects with theophylline, if so which ones?*

*13) a. Was your child treated in the past with any of these medications: Acetazolamide, Caffeine,* ***Clonazepam****,* ***Diazepam****, Levetiracetam,* ***Levodopa****, Tetrabenazine, Trihexyphenidyl, or others?*

*b. Is your child still being treated with one or more of these medications? Please specify which one(s).*

*c. On a scale of 0 (no improvement) to 10 (major improvement with total disappearance of symptoms), how would you rate the improvement with this medication on his/her involuntary movements?*

0 1 2 3 4 5 6 7 8 9 10
